# Joint Effects of Indoor Air Pollution and Maternal Psychosocial Factors During Pregnancy on Trajectories of Early Childhood Psychopathology

**DOI:** 10.1101/2023.04.07.23288289

**Authors:** Grace M. Christensen, Michele Marcus, Aneesa Vanker, Stephanie M. Eick, Susan Malcolm-Smith, Shakira F. Suglia, Howard H. Chang, Heather J. Zar, Dan J. Stein, Anke Hüls

**Affiliations:** Department of Epidemiology, Rollins School of Public Health, Emory University, Atlanta, GA, USA; Gangarosa Department of Environmental Health, Rollins School of Public Health, Emory University, Atlanta, GA, USA; Department of Paediatrics and Child Health, Red Cross War Memorial Children’s Hospital, University of Cape Town, Cape Town, South Africa; Neuroscience Institute, University of Cape Town, Cape Town, South Africa; Department of Psychiatry and Mental Health, University of Cape Town, Cape Town, South Africa; Department of Biostatistics, Rollins School of Public Health, Emory University, Atlanta, GA, USA; South African Medical Research Council (SAMRC) Unit on Risk and Resilience in Mental Disorders, University of Cape Town, Cape Town, South Africa

## Abstract

**Background:** Prenatal indoor air pollution and maternal psychosocial factors have been associated with adverse psychopathology. We used environmental exposure mixture methodology to investigate joint effects of both exposure classes on child behavior trajectories.

**Methods:** For 360 children from the South African Drakenstein Child Health Study, we created trajectories of Child Behavior Checklist scores (24, 42, 60 months) using latent class linear mixed effects models. Indoor air pollutants and psychosocial factors were measured during pregnancy (2^nd^ trimester). After adjusting for confounding, single-exposure effects (per natural log-1 unit increase) were assessed using polytomous logistic regression models; joint effects using self-organizing maps (SOM), and principal component (PC) analysis.

**Results:** High externalizing trajectory was associated with increased particulate matter (PM_10_) exposure (OR [95%-CI]: 1.25 [1.01,1.55]) and SOM exposure profile most associated with smoking (2.67 [1.14,6.27]). Medium internalizing trajectory was associated with increased emotional intimate partner violence (2.66 [1.17,5.57]), increasing trajectory with increased benzene (1.24 [1.02,1.51]) and toluene (1.21 [1.02,1.44]) and the PC most correlated with benzene and toluene (1.25 [1.02, 1.54]).

**Conclusions:** Prenatal exposure to environmental pollutants and psychosocial factors was associated with internalizing and externalizing child behavior trajectories. Understanding joint effects of adverse exposure mixtures will facilitate targeted interventions to prevent childhood psychopathology.

## Introduction

Childhood psychopathology, including emotional and behavioral problems, affect parents, teachers, and most importantly the child, and can persist into adulthood^1^. Experiencing mental health problems before age 14 is associated with increased risk of adult psychopathology.

Psychopathology is characteristically split into two categories of disorders, internalizing and externalizing. Externalizing behaviors reflect the behavior towards the environment, whereas internalizing behaviors are reflected inward. Externalizing conditions include attention-deficit/hyperactivity disorder (ADHD) and oppositional defiant, and internalizing conditions include anxiety and depression^2,3^.

Pregnancy is a sensitive period of brain development, as the central nervous system begins to develop as early as the first month of gestation^4^. Investigating modifiable risk factors of childhood psychopathology during pregnancy has the potential to reduce the burden of mental health problems in both children and adults^5^.

Indoor air pollution is a ubiquitous and well-known contributor to the global burden of disease ^6,7^. Animal studies have shown exposure to air pollutants during pregnancy affects the central nervous system of the fetus and elements of behavior in adulthood^8,9^. Epidemiologic studies in humans have also shown that exposure to outdoor air pollutants during pregnancy and early life affects child psychopathology^10–12^. A Korean study found maternal smoking during pregnancy was associated with adverse Childhood Behavior Checklist (CBCL) total problems score at 5 years^13^. Additionally, indoor air pollutants from maternal cooking during pregnancy associated with hyperactive behaviors in children at 3 years old, though the study did not measure individual air pollutants, and instead used survey information on cooking fuel as a proxy measure^14^.

Exposure to adverse psychosocial factors during pregnancy also negatively affects child development and mental health. Animal studies have shown prenatal stress affects behavior^15–17^, and there is growing epidemiological evidence that prenatal psychosocial stressors are associated with impaired child cognitive development^18^. The Australian Raine study found stressful life events during pregnancy, family income below poverty line, and mother not finishing high school were associated with increased CBCL t-scores from age 2-14 years^19^.

Additionally, stressful events during pregnancy, including death of a relative, and financial problems, was associated with increased levels of behavior problems in childhood and problematic mental health trajectories in the Raine study^19,20^.

Childhood psychopathology can be impacted by both psychosocial and environmental factors, and joint effects of these exposures are poorly understood^21^, as they are usually researched separately. Synergistic effects of environmental and psychosocial factors are probable, and have been demonstrated in previous research on birth outcomes like birthweight and gestational age^21–24^. It is important to investigate joint and potentially synergistic effects of environmental and psychosocial factors because it may help to identify especially vulnerable subgroups, in which the disease burden is likely to be larger than in the general population. So far, very few studies have investigated joint effects of air pollution and psychosocial factors on childhood psychopathology. One epidemiologic study conducted in New York City, USA, found that prenatal exposure to polyaromatic hydrocarbons increased the effect of exposure to psychosocial stress on CBCL score in school-age children ^11^. Another study, conducted in Boston, USA, found Black Carbon exposure during pregnancy was significantly associated with lower attention concentration index scores in boys with high exposure to prenatal stress^25^.

However, both of these studies only used one air pollutant and one psychosocial factor when investigating joint effects of air pollution and psychosocial stress on child psychology instead of an exposure mixture. This does not reflect real life exposure as people are exposed to many pollutants and psychosocial factors at the same time^21^. Additionally, both of these studies were conducted in the USA, a high-income country.

A majority of research on childhood psychopathology is conducted in high income countries, yet almost 80% of children live in low to middle income countries (LMICs)^26^. Indoor air pollution is an important source of air pollution in LMICs, where many homes rely on alternate fuel sources for household energy and particularly affects women who through traditional gender norms generally spend more time indoors and are more involved in food preparation and cooking^6,27^.

Additionally, in LMICs, prevalence of perinatal depression and exposure to violence is even higher than in high income countires^28^. Therefore, pregnant women in LMICs may be uniquely susceptible to the joint effects of indoor air pollution and psychosocial factors.

We aim to investigate the individual and joint effects of prenatal exposure to indoor air pollution and maternal psychosocial factors on trajectories of psychopathology in early childhood in a South African birth cohort. This study uses traditional single-exposure polytomous logistic regression modeling to investigate the effects of indoor air pollutants and psychosocial factors during pregnancy individually, as well as exposure mixture methods, such as self-organizing maps and principal components analysis, to investigate joint effects of exposures.

## Methods

### Data Source

This study uses data from a subset of participants enrolled in the Drakenstein Child Health Study (DCHS), a multi-disciplinary population-based pregnancy cohort based in South Africa. Pregnant women were recruited in their 2^nd^ trimester of pregnancy from 2012-2015, follow-up with mother-child pairs has been conducted annually thereafter. Pregnant women seeking care at two public sector primary healthcare clinics, who were at least 18 years of age, were within 20-28 weeks gestation, and had no intention of moving away from the district were eligible for enrollment. Recruitment for DCHS has been described elsewhere^28,29^. The full cohort includes N=1,141 mother-child pairs, among which a subset (n=819) were selected for indoor air pollution measurement. Mother-child pairs with indoor air pollution measurements, and complete child behavior checklist measurements at 24, 42, and 60 months of age were included in this analysis (n=360). The DCHS was approved by the Human Research Ethics Committee of the Faculty of Health Sciences, University of Cape Town, by Stellenbosch University and the Western Cape Provincial Research committee. Written informed consent was provided by the mothers for herself and her child and is renewed annually.

### Indoor Air Pollution Assessment

Indoor air pollution measurements were taken during participants’ 2^nd^ trimester of pregnancy. Pollutants measured include particulate matter <10 microns in diameter (PM_10_), carbon monoxide (CO), nitrogen dioxide (NO_2_), sulfur dioxide (SO_2_), and Volatile Organic Compounds (VOCs) benzene and toluene. Particulate Matter (PM_10_) was collected over 24 hours with a personal air sampling pump (SKC AirChek 52®), using a gravimetrically pre-weighted filter.

Carbon monoxide (CO) was collected over 24 hours using an Altair® carbon monoxide single gas detection unit, electrochemical sensor detection of gas at 10-minute intervals were collected. Sulphur dioxide (SO_2_) and nitrogen dioxide (NO_2_) were collected over 2 weeks using Radiello® absorbent filters in polyethylene diffusive body. Volatile organic compounds, including benzene and toluene, were collected over 2 weeks using Markes® thermal desorption tubes^30^. Information on type of home, distance from major road, size of home, number of inhabitants, access to basic amenities, fuels used for cooking and heating, ventilation within homes, and pesticides and cleaning materials used in the home was collected at home visits^30^.

### Assessment of Psychosocial Factors

Psychosocial factors were collected via questionnaire in the 2^nd^ trimester of pregnancy and included multiple dimensions of psychosocial stress. Employment, education, household income, household assets, marital status, number of dependents, and financial activities were included as indicators of socioeconomic status. Perceived household food insecurity was assessed using an adapted version of the USDA Household Food Security Scale^31^. Intimate partner violence was assessed using the IPV Questionnaire adapted from the WHO multi-country study and the Women’s Health Study in Zimbabwe^32,33^. The IPV questionnaire assesses lifetime and recent (past year) exposure to emotional, physical, and sexual violence. The World Mental Health Life Events Questionnaire (LEQ) was used to measure trauma and resilience.

Use of alcohol and tobacco were assessed using the Alcohol, Smoking, and Substance Involvement Screening Test (ASSIST). Additionally, tobacco smoke exposure was assessed via urinary cotinine and questionnaire. The Self Reporting Questionnaire (SRQ-20), a measure endorsed by the WHO, was used to measure psychological distress^34,35^. The Edinburgh Postnatal Depression Scale (EPDS) was used to measure depressive symptoms^36^.

### Outcome Assessment

Parent-reported child psychopathology was assessed using the pre-school version of the Child Behavior Checklist (CBCL) administered at 24, 42, and 60 months old^37^. Child behavior was assessed using a 3-point Likert scale (0 = not true; 2 = often or very true) to create a score, consisting of 113 questions, which can be divided into internalizing and externalizing sub scores. The internalizing scale combines the scores from the anxious/depressed, withdrawn/depressed, and somatic complaints syndromic scales. The externalizing scale combines the rule-breaking and aggressive behavior syndromic scales^37^. Higher CBCL scores indicate increased problematic behavior, indicative of child psychopathology. CBCL scores show good associations with psychopathology diagnoses from the DSM-5 including, anxiety, oppositional defiant disorder, attention deficit/hyperactivity disorder, among others^38^.

Standardized CBCL scores, or T-scores, for externalizing and internalizing behavior were used as outcomes in our analyses.

### Statistical Analysis

#### Multiple imputation of missing values

While there were no missing values in any of the outcome variables or covariates in our final analysis sample, some participants are missing indoor air pollution or psychosocial measurements (Table S2). We assume these missing exposure variables are missing at random based on inspections of missingness patterns (Figure S1). To increase the sample size, we used multiple imputation to impute these missing exposure values. Using the R package *Hmisc*, indoor air pollution and psychosocial factor variables were imputed using predictive mean matching, with models that include indoor air pollutants, house characteristics, and psychosocial factor measures. Five seed numbers were created using a random number generator, each seed resulted in its own set of multiple imputed variables. One imputed set, from the 5 sets, from each seed was randomly chosen to use for analyses. The seed with the highest R^2^ values, a measure used to explain how well the missing variable was predicted, was selected for primary analysis. R^2^ values for each multiple imputed exposure variable differed between seeds. Analyses using complete cases and the other imputation seeds were conducted as a sensitivity analysis.

#### Assessment of CBCL Trajectories

The outcome used in this study was trajectory of CBCL score from 24, 42, and 60 months old children. Participants with CBCL measurements at all time points (n=360) were included.

Trajectories were created using Latent Class Linear Mixed Effects Models (LCMM). LCMM models for log-transformed CBCL t-scores were used to create latent classes, using child sex as a fixed effect covariate and age in months at CBCL measurement as a random effect covariate. Using the R package *lcmm*, LCMM defines a number of ‘typical’ trajectories of CBCL scores, which then were assigned to participants and used as the outcome in the subsequent polytomous logistic regression analyses (described below)^39^. One to five latent classes were evaluated in LCMM models, the final model was selected based on measures of model fit such as Akaike Information Criterion (AIC), Bayesian Information Criterion (BIC), sample-size-adjusted BIC (SABIC), and entropy (Table S3). Trajectories for CBCL externalizing and internalizing sub scales were created.

#### Single-exposure models

Adjusted polytomous logistic regression models were used to estimate single air pollutant and psychosocial factor effects on CBCL trajectory. Confounding was assessed using directed acyclic graphs (DAGs) informed by prior research and literature reviews (Figure S2). To control for confounding, each model was adjusted for maternal age at baseline, maternal HIV status, child ancestry, and socioeconomic status (when not used as the exposure of interest). In individual models, all exposures were natural log-transformed for modeling. To account for confounding by psychosocial factors, models with indoor air pollutants as the main exposure were additionally adjusted for psychosocial factors, and vice versa. The model estimating the effect of socioeconomic status was adjusted air pollution exposures. To avoid oversaturation of the linear regression models, and as the exposures within each exposure group (air pollution exposure and psychosocial factors) were highly correlated, the first principal components of each group were used as confounders instead of the original variables. To create these principal components, we conducted a principal component analysis (PCA) for each exposure group separately (more details can be found in the supplementary methods). In a sensitivity analysis to account for seasonality of indoor air pollution measurement, we adjusted the air pollution exposure models for season of indoor air pollution measurement along with previously mentioned confounders.

#### Joint effects models

We used two complementary mixture methods to examine joint effects of indoor air pollution and psychosocial factors: PCA and self-organizing maps (SOM). While PCA was not developed as an exposure mixture method, using the first few PCs explains a percentage of the total variance in the data using a smaller number of variables. In addition, PCs are continuous, orthogonal and uncorrelated, which can increase the statistical power to detect associations in comparison to categorical variables. PCs were calculated based on centered and scaled indoor air pollution and psychosocial exposure variables. PCs of the exposure mixture (combination of indoor air pollution and psychosocial exposure variables) were created and added as exposure variables to the polytomous logistic regression model to investigate the joint effect of indoor air pollution and psychosocial factors on CBCL trajectories. The number of PCs added to the model was determined by proportion of variance contributed as seen in an ‘elbow plot’ (Figure S3), resulting in five PCs that explained 55% of the total variance. Polytomous logistic regression models were adjusted for maternal age at baseline, maternal HIV status, and child ancestry.

SOM was used to examine the effects of certain exposure profiles on CBCL trajectory. The SOM algorithm identifies exposure cluster profiles with exposure levels homogenous within the cluster and heterogeneous between clusters. The advantage of SOM over PCA is interpretability of the exposure profile clusters. However, with smaller sample sizes there can be low numbers of participants in some clusters. The number of clusters chosen for analyses was based on multiple statistical measures identifying group structure, including AIC, and adjusted R^2^, as well as visual inspection of the clusters for interpretability and suitable number of participants in each cluster, resulting in four SOM clusters to represent prenatal exposure profiles. Indoor air pollution and psychosocial factor variables were centered and scaled before running the SOM clustering algorithm. The effect of these exposure clusters on CBCL trajectory was assessed using adjusted polytomous logistic regression. Adjusted models were adjusted for maternal age, maternal HIV status, and child ancestry. We used the SOM R package as implemented in https://github.com/johnlpearce/ECM.

All analyses were performed using R version 3.6.1 (R Core Team, Vienna, Austria).

## Results

### Study Population

The final sample for this analysis consisted of 360 mother-child pairs. The mean maternal age during pregnancy was 26.9 (SD = 5.6) years. Nearly a quarter of mothers (n=78; 21.7%) were HIV positive at baseline. Half of the children were male (n=190; 52.8%). In this sample 50.3% (n=181) of mothers identified their children as having mixed ancestry, the other half identified as having black African ancestry (n=179; 49.7%) (Table 1). Table S1 compares demographic and exposure characteristics in the full cohort, the indoor air pollution subsample, and the analysis sample. Demographic characteristics and psychosocial factor scores are similar across samples. Indoor air pollutant exposure concentrations in the analysis sample are slightly lower than in the full cohort, except for PM_10_ (analysis sample median: 41.46 µg/m^3^ vs full cohort:

**Table 1.**
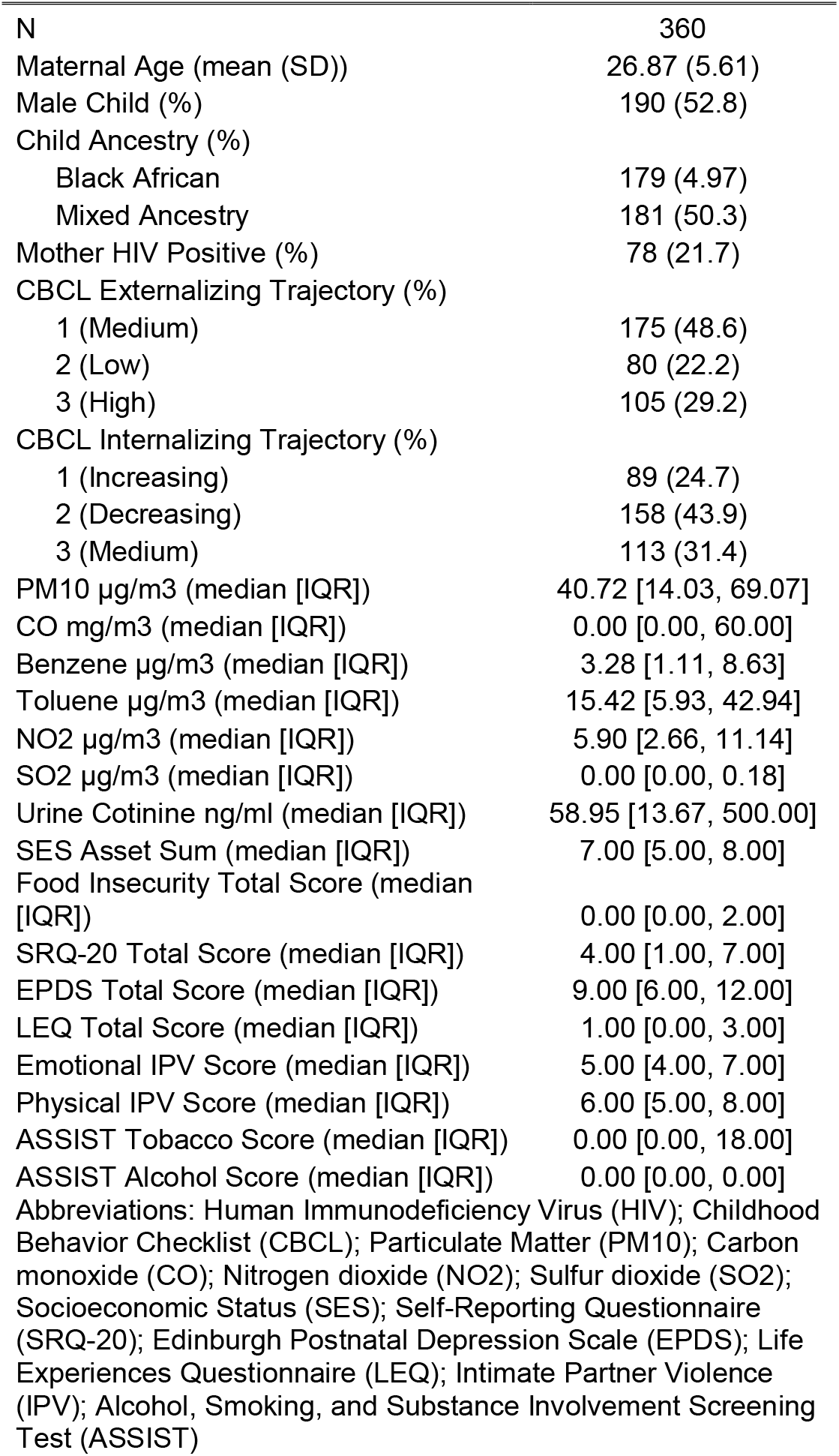
Drakenstein Child Health Study (DCHS) population characteristics.

|  |  |
| --- | --- |
| N | 360 |
| Maternal Age (mean (SD)) | 26.87 (5.61) |
| Male Child (%) | 190 (52.8) |
| Child Ancestry (%) |  |
| Black African | 179 (4.97) |
| Mixed Ancestry | 181 (50.3) |
| Mother HIV Positive (%) | 78 (21.7) |
| CBCL Externalizing Trajectory (%) |  |
| 1 (Medium) | 175 (48.6) |
| 2 (Low) | 80 (22.2) |
| 3 (High) | 105 (29.2) |
| CBCL Internalizing Trajectory (%) |  |
| 1 (Increasing) | 89 (24.7) |
| 2 (Decreasing) | 158 (43.9) |
| 3 (Medium) | 113 (31.4) |
| PM10 µg/m3 (median [IQR]) | 40.72 [14.03, 69.07] |
| CO mg/m3 (median [IQR]) | 0.00 [0.00, 60.00] |
| Benzene µg/m3 (median [IQR]) | 3.28 [1.11, 8.63] |
| Toluene µg/m3 (median [IQR]) | 15.42 [5.93, 42.94] |
| NO2 µg/m3 (median [IQR]) | 5.90 [2.66, 11.14] |
| SO2 µg/m3 (median [IQR]) | 0.00 [0.00, 0.18] |
| Urine Cotinine ng/ml (median [IQR]) | 58.95 [13.67, 500.00] |
| SES Asset Sum (median [IQR]) | 7.00 [5.00, 8.00] |
| Food Insecurity Total Score (median [IQR]) | 0.00 [0.00, 2.00] |
| SRQ-20 Total Score (median [IQR]) | 4.00 [1.00, 7.00] |
| EPDS Total Score (median [IQR]) | 9.00 [6.00, 12.00] |
| LEQ Total Score (median [IQR]) | 1.00 [0.00, 3.00] |
| Emotional IPV Score (median [IQR]) | 5.00 [4.00, 7.00] |
| Physical IPV Score (median [IQR]) | 6.00 [5.00, 8.00] |
| ASSIST Tobacco Score (median [IQR]) | 0.00 [0.00, 18.00] |
| ASSIST Alcohol Score (median [IQR]) | 0.00 [0.00, 0.00] |
| Abbreviations: Human Immunodeficiency Virus (HIV); Childhood Behavior Checklist (CBCL); Particulate Matter (PM10); Carbon monoxide (CO); Nitrogen dioxide (NO2); Sulfur dioxide (SO2); Socioeconomic Status (SES); Self-Reporting Questionnaire (SRQ-20); Edinburgh Postnatal Depression Scale (EPDS); Life Experiences Questionnaire (LEQ); Intimate Partner Violence (IPV); Alcohol, Smoking, and Substance Involvement Screening Test (ASSIST) |  |

33.45 µg/m^3^) and cotinine (analysis sample median: 61.34 ng/mL vs full cohort: 43.00 ng/mL) which are higher than in the full cohort.

### Trajectories

At each time period, CBCL internalizing and externalizing sub scores were highly correlated (24 months: Pearson r = 0.71; 42 months: r = 0.7; 60 months: r = 0.82; Table S5).

Three trajectories were chosen for CBCL externalizing problems and internalizing problems (Table S3). Trajectories for externalizing problems were categorized as ‘high’, ‘medium’, and ‘low’ trajectories, as they correspond to relatively high, medium, and low scores across time points (Figure 1A, Table S6). Latent class 1, the ‘medium’ trajectory decreased slightly over time but scores were always between the ‘high’ and ‘low’ trajectory. Latent class 2 had the lowest CBCL externalizing scores over the study period. In contrast, latent class 3 always had the highest CBCL externalizing score over the study period. In all regression analyses the ‘low’ trajectory was used as the reference group.

**Figure 1.**
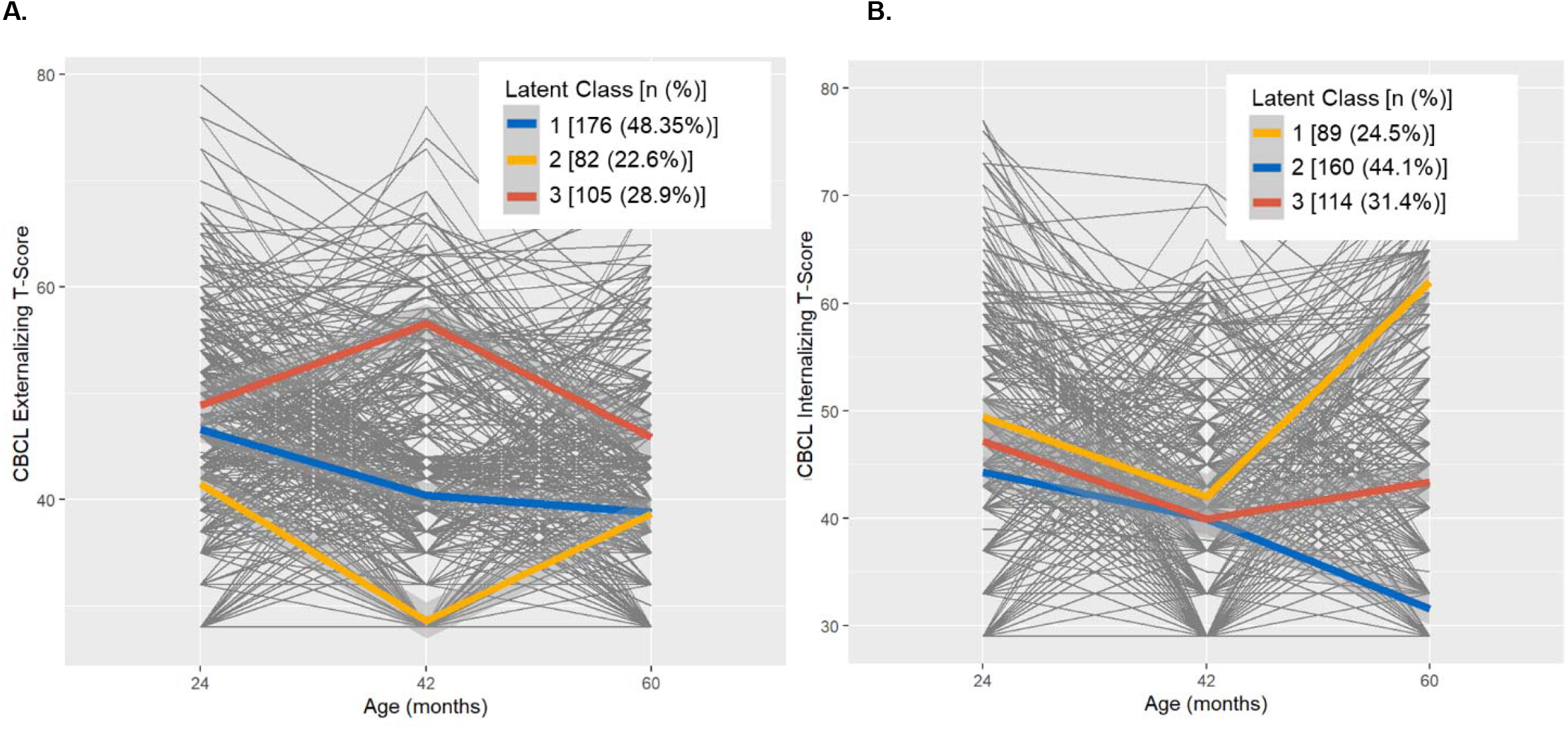
Latent Class Mixed Model (LCMM) trajectories. Child Behavior Checklist (CBCL) T-score trajectories modeled using LCMM, adjusted for child sex and age in months at CBCL assessment in the DCHS. **A**. CBCL externalizing problems T-score. **B**. CBCL Internalizing problems T score.

CBCL internalizing problems score trajectories were categorized as, having ‘decreasing’, ‘medium’ and ‘increasing’ trajectories., For the internalizing problems scores, all three trajectories slightly decreased between 24 and 42 months and diverged from 42 to 60 months (Figure 1B, Table S6). The CBCL internalizing problems trajectory shown in latent class 1 is characterized as having sharply increasing CBCL internalizing score after 42 months, this trajectory is described as the ‘increasing’ trajectory. In contrast, latent class 2 shows a decreasing trajectory over the time period and will be described as the ‘decreasing’ trajectory. Latent class 3 is stable over time and scores are between the increasing and decreasing trajectories, therefore will be described as the ‘medium’ trajectory. In all regression analyses the ‘decreasing’ trajectory was used as the reference group.

### Single-exposure models

CBCL externalizing problems trajectories showed an adverse association with PM_10_, which was associated with the high trajectory (1.25; 1.01, 1.55) (Figure 2A, Table S7). CBCL internalizing problems showed an adverse association with two air pollutants (benzene and toluene) and one psychosocial factor (emotional IPV score). Increasing internalizing CBCL trajectory was associated with both increased log-transformed benzene (1.24; 1.02, 1.51) and toluene (1.21; 1.02, 1.44) exposure. Emotional IPV score was associated with higher odds of the medium trajectory (2.66; 1.27, 5.57) (Figure 2B, Table S7).

**Figure 2.**
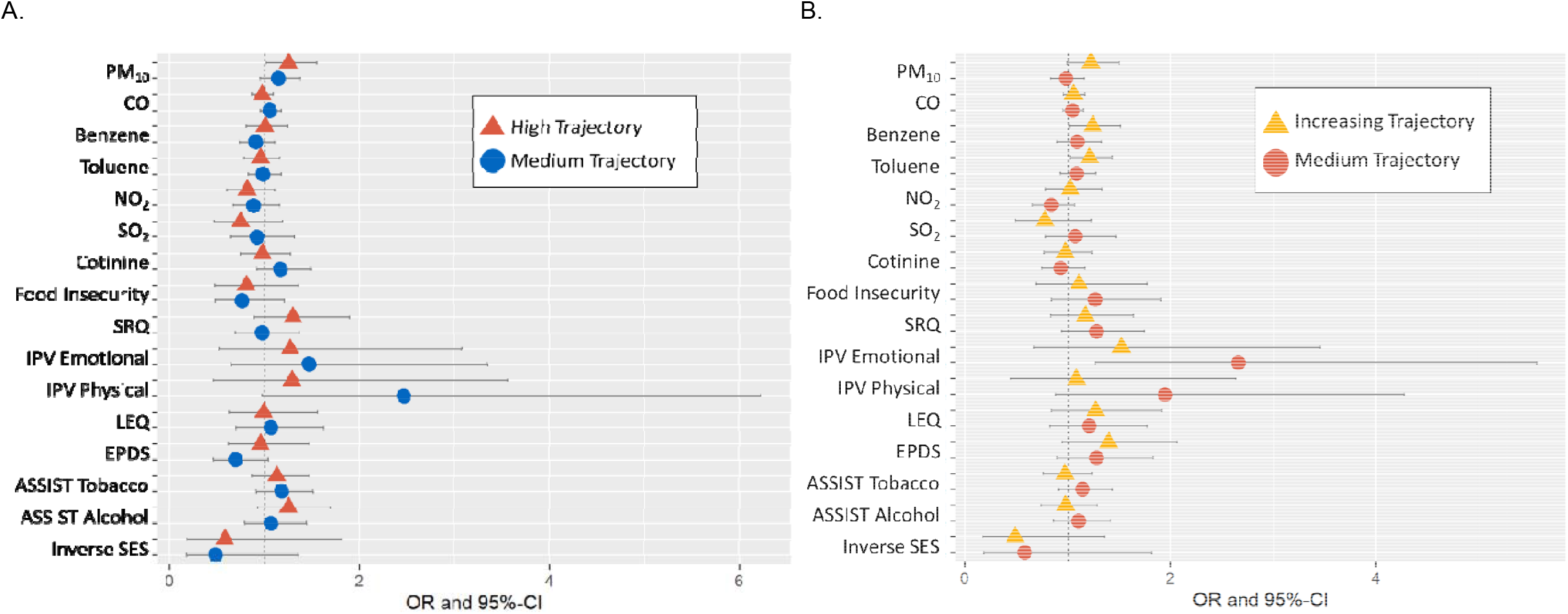
Results from single-exposure polytomous logistic regression models, adjusted for maternal age, maternal HIV status, and ancestry, in the DCHS (N=360). A. CBCL Externalizing Problems. B. CBCL Internalizing Problems.

### Joint effects models

In adjusted polytomous logistic regression models using externalizing CBCL trajectory, PC1 was associated with both high (1.25; 1.02, 1.54) and medium (1.27; 1.04, 1.54) trajectories and PC3 was significantly associated with the medium (1.33; 1.04, 171) trajectory compared to the low trajectory. Both PC1 and 3 are explained by high cotinine level and high ASSIST Tobacco and Alcohol scores. The more robust association with PC1 reflects that externalizing CBCL trajectory is associated with both smoking related exposures and high psychosocial stressors (Figure 3A, Table S8). Increasing internalizing CBCL trajectory was associated with PC2 (1.22; 1.02, 1.48), the PC mostly explained by high benzene and toluene levels in adjusted polytomous regression models (Figure 3B, Table S8).

**Figure 3.**
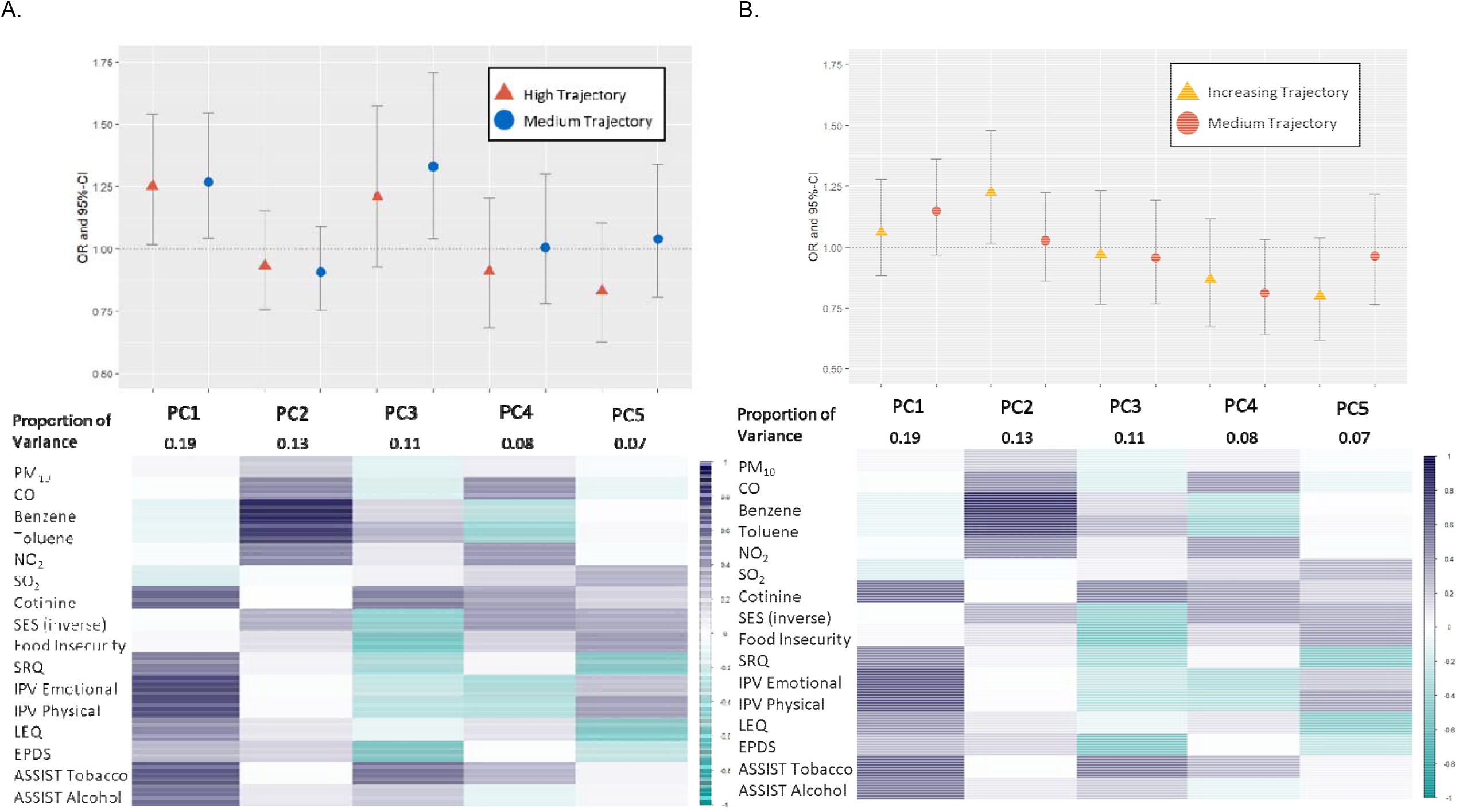
Top panels show results of polytomous logistic regression modeling using principal components of exposure mixture, adjusted for maternal age, maternal HIV status, ancestry in the DCHS. Bottom panels show a correlation matrix between individual air pollutant and psychosocial factor exposures and each principal component. Purple indicates higher positive correlation, while teal indicated higher negative correlation in the DCHS. A. CBCL Externalizing Problems. B. CBCL Internalizing Problems.

In SOM analyses with externalizing problems trajectory, the cluster associated with high cotinine level and ASSIST tobacco and alcohol scores (cluster 3) was associated with the high trajectory (2.67; 1.14, 6.27), compared to the low exposure cluster (cluster 1) (Figure S4, Table S9), in line with the PCA analysis. No SOM exposure cluster was associated with CBCL internalizing problems trajectories.

### Sensitivity analyses

Results from sensitivity analysis models comparing complete case and imputed exposures were similar to the main results presented above. In internalizing problems models, toluene was also associated with the increasing trajectory in one other imputation model, but ORs were consistent across all models. Similarly for benzene, the association with increasing trajectory was not significant for other models, but the ORs consistent across all models (Table S7).

## Discussion

In this analysis of mother-child pairs from a South African birth cohort, trajectories of CBCL T-scores at 24, 42, and 60 months were differentially associated with prenatal indoor air pollution and psychosocial exposures. CBCL internalizing problems trajectory was individually associated with both exposures to indoor air pollution and adverse psychosocial factors, while the externalizing problems trajectory was mostly associated with smoking related exposures. This analysis indicates that different exposures might differently affect internalizing and externalizing problem trajectories.

We observed that externalizing problems trajectory was most associated with prenatal smoking behaviors and PM_10_, a by-product of cigarette smoke. This has been observed in a previous study from the DCHS^40^. In other prior studies investigating PM_10_ exposure during pregnancy and autism spectrum disorder, a condition that exhibits externalizing behaviors, results have been mixed. Only one study found a significant harmful effect of PM_10_^41^, the rest found a null association^42^. Other studies investigating smoking during pregnancy have found associations with externalizing behaviors, including inattention and impulsivity^43–45^.

The internalizing problems trajectory was associated with emotional IPV. Few studies have investigated the association between IPV before and during pregnancy and childhood psychopathology. One study found IPV during pregnancy was correlated with borderline or clinical internalizing problems, externalizing problems and total problems using the CBCL in children 18 months-18 years old^46^. However, results from our study may not be directly comparable as that study did not separate different types of IPV, and did not adjust for confounding. More epidemiology studies investigating prenatal exposure to IPV and their association with child behavior are needed. Our study also found individual and joint associations with VOCs, specifically benzene and toluene, and internalizing problems trajectory.

To our knowledge, our study is the first to investigate the association between prenatal VOC exposure and childhood psychopathology. One prior study has investigated VOCs and neurodevelopmental outcomes, though they measured exposure in early childhood, and found VOCs m,p-xylene and o-xylene were associated with decreased scores in the ages and stages questionnaire, which screens young children for developmental delays^47^. Continued epidemiologic research on prenatal VOC exposure is necessary to examine the relationship between VOCs and child psychopathy.

In support of our findings, prior studies investigating neurodevelopmental or psychopathological outcomes at one time period found an interaction between air pollutants and adverse psychosocial factors. However, these studies did not use environmental mixture methods and instead used interaction terms between one air pollutant and one psychosocial factor^11,25,48,49^.

Studies using other health outcomes have also found joint effects of prenatal air pollution and psychosocial factor exposure. A recent review article found several studies showing prenatal psychosocial factors modifying the effect of ambient and traffic related air pollution on adverse birth and childhood outcomes such a birthweight, gestational age and asthma^21^.

Synergy is probable because air pollutants and stress caused by psychosocial factors may impact similar brain mechanisms, including inflammation ^9,50^. It is hypothesized that air pollution affects the central nervous system (CNS) via neuroinflammation and oxidative stress^9,51–54^.

Animal models show that air pollutants cause a systemic inflammatory response, including neuroinflammation inside the brain. Both the physical air pollutant particle, and the toxic components absorbed on the particle can create an inflammatory response. Translocation of air pollutant nanoparticles from the lungs and nasal pathways to other areas of the body, including the placenta^55^, cause damage to the mother’s body and fetus. Microglia respond to this damage by releasing inflammatory cytokines like TNF-α, IL-1β, and IL-6, and reactive oxygen species (ROS), inducing oxidative stress. Chronic activation of the microglia and over production of inflammatory markers and ROS can cause neuronal cell death^9^. A range of research also documents that adverse psychosocial factors are associated with neuroinflammation and oxidative stress^50,54,56^.

Several strengths of this study deserve emphasis. First, this analysis used measurements of multiple indoor air pollution exposures. Indoor air pollution in LMIC is most often measured using survey-based proxy measures e.g. cooking practices, smoking, etc. Given the well-known harmful effects of indoor air pollution^27^, it is important to measure individual pollutants to pinpoint which chemical, or combination of chemicals, is causing health problems and the biological mechanisms involved. Second, this study leverages a unique prospective birth cohort with repeated CBCL measurements across time periods. And finally, the traditional single-exposure analysis is complemented with two environmental mixtures methods, PCA and SOM. These methods allow us to explore joint-effects of environmental and social exposure profiles that are associated with psychopathology trajectory. Estimating joint effects can identify vulnerable subgroups to target for interventions to reduce childhood psychopathology.

Several limitations deserve emphasis. First, we were unable to investigate the effect of the total mixture on CBCL trajectories. Currently, environmental mixture methods that estimate a total mixture effect (e.g. Bayesian kernel machine regression, weighted quantile sum regression, etc.) can only accommodate linear or logistic regression, and not polytomous logistic regression. As methods to investigate environmental exposure mixtures advance this may be an option for future studies. Second, the lack of fine (PM_2.5_) and ultrafine PM measurements. PM_2.5_ measurement was not collected because, at the time, personal PM_2.5_ monitoring was not easily available for this large of a study. Additionally, this study uses indoor air pollution measurements from one point (24 hours or two-week average, depending on pollutant) in the 2^nd^ trimester and in early life to characterize exposure for the whole period. This could create some misclassification of the exposure. Though prior research in the DCHS has found associations between these exposures and several outcomes^57,58^, including neurodevelopment^59^. Other limitations include biases from residual confounding, and selection bias, though this cohort was selected to be population based and representative of peri-urban populations in South Africa and other low income countries. While this study had a relatively small sample size, few studies have such detailed exposure information and repeated outcomes, especially in a low to middle income country.

## Conclusion

We observed single-exposure and joint effects of prenatal exposure to indoor air pollutants and psychosocial factors on trajectories of childhood psychopathology in 24-60 months old children from South Africa. Externalizing and internalizing trajectories were associated with prenatal indoor air pollution and psychosocial factors. Investigating joint effects of environmental and social exposures is necessary for identifying vulnerable subgroups to target for interventions to reduce exposure and burden of childhood psychopathology.

## Data Availability

All data produced in the present study are available upon reasonable request to the authors

