## Supplemental Materials for "Joint Effects of Indoor Air Pollution and Maternal Psychosocial Factors During Pregnancy on Trajectories of Early Childhood Psychopathology"

**Supplementary Methods**

Principal Components Analysis (PCA) use in single-exposure modeling for covariate adjustment

To avoid oversaturation of the linear regression models, and as the exposures within each exposure group (air pollution exposure and psychosocial factors) were highly correlated, the first principal components of each group were used as confounders instead of the original variables. To create these principal components, we conducted a principal component analysis (PCA) for each exposure group separately and included the principal components (PC) as covariates in the models to adjust for psychosocial factors or air pollution exposure. PCA is a dimension reduction approach that creates PCs as linear combinations of input variables. The number of PCs added to the model was determined by proportion of variance contributed as seen in an ‘elbow plot’, resulting in 3 PCs that explained 56% of the total variance of psychosocial factors and 3 PCs that explained 58% of the total variance of air pollution exposure.

**Supplemental Tables**

**Table S1.** Comparison of demographic and exposure characteristics between the full DCHS cohort, the subsample with indoor air pollution measurements, and the analysis sample.

|  | Full DCHS Cohort | IAP Subsample | Analysis Sample |
| --- | --- | --- | --- |
| n | 1143 | 819 | 360 |
| Maternal Age (mean (SD)) | 26.60 (5.68) | 26.60 (5.67) | 26.87 (5.61) |
| Male Child (%) | 586 (51.3) | 422 (51.5) | 190 (52.8) |
| Mixed Ancestry (%) | 510 (44.7) | 379 (46.3) | 181 (50.3) |
| Mother HIV Positive (%) | 248 (21.7) | 171 (20.9) | 78 (21.7) |
| PM10 µg/m3 (median [IQR]) | 33.37 [12.49, 64.80] | 33.45 [12.49, 65.43] | 41.46 [14.42, 69.71] |
| CO mg/m3 (median [IQR]) | 0.00 [0.00, 102.50] | 0.00 [0.00, 120.00] | 0.00 [0.00, 0.00] |
| Benzene µg/m3 (median [IQR]) | 4.28 [1.75, 11.29] | 4.34 [1.75, 11.50] | 3.21 [1.05, 8.47] |
| Toluene µg/m3 (median [IQR]) | 16.79 [7.04, 44.24] | 16.94 [7.09, 44.79] | 15.51 [5.80, 44.47] |
| NO2 µg/m3 (median [IQR]) | 7.13 [3.33, 12.69] | 7.19 [3.34, 12.70] | 5.75 [2.63, 11.21] |
| SO2 µg/m3 (median [IQR]) | 0.00 [0.00, 0.28] | 0.00 [0.00, 0.28] | 0.00 [0.00, 0.17] |
| Urine Cotinine ng/ml (median [IQR]) | 43.00 [10.70, 500.00] | 43.35 [10.70, 500.00] | 61.35 [14.33, 500.00] |
| SES Asset Sum (median [IQR]) | 7.00 [5.00, 8.00] | 7.00 [5.00, 8.00] | 7.00 [5.00, 8.00] |
| Food Insecurity Total Score (median [IQR]) | 0.00 [0.00, 2.00] | 0.00 [0.00, 2.00] | 0.00 [0.00, 2.00] |
| SRQ-20 Total Score (median [IQR]) | 4.00 [1.00, 7.00] | 4.00 [1.50, 7.00] | 3.00 [1.00, 6.00] |
| EPDS Total Score (median [IQR]) | 9.00 [6.00, 12.00] | 9.00 [6.00, 13.00] | 9.00 [6.00, 12.00] |
| LEQ Total Score (median [IQR]) | 1.00 [0.00, 3.00] | 1.00 [0.00, 3.00] | 1.00 [0.00, 3.00] |
| Emotional IPV Score (median [IQR]) | 4.00 [4.00, 7.00] | 5.00 [4.00, 7.00] | 5.00 [4.00, 7.00] |
| Physical IPV Score (median [IQR]) | 5.00 [5.00, 7.00] | 5.00 [5.00, 7.00] | 6.00 [5.00, 8.00] |
| ASSIST Tobacco Score (median [IQR]) | 0.00 [0.00, 13.00] | 0.00 [0.00, 14.00] | 0.00 [0.00, 18.00] |
| ASSIST Alcohol Score (median [IQR]) | 0.00 [0.00, 0.00] | 0.00 [0.00, 0.00] | 0.00 [0.00, 0.00] |
| Abbreviations: Indoor Air Pollution (IAP); Human Immunodeficiency Virus (HIV); Particulate Matter (PM10); Carbon monoxide (CO); Nitrogen dioxide (NO2); Sulfur dioxide (SO2); Socioeconomic Status (SES); Self-Reporting Questionnaire (SRQ-20); Edinburgh Postnatal Depression Scale (EPDS); Life Experiences Questionnaire (LEQ); Intimate Partner Violence (IPV); Alcohol, Smoking, and Substance Involvement Screening Test (ASSIST) | | | |

**Table S2.** Proportion of missing prenatal exposure data in analysis sample.

| **Exposure** | **# missing** | **% missing** | **n** | **total N*** |
| --- | --- | --- | --- | --- |
| PM10 | 38 | 11% | 322 | 360 |
| CO | 74 | 21% | 286 | 360 |
| benzene | 49 | 14% | 311 | 360 |
| toluene | 49 | 14% | 311 | 360 |
| NO2 | 43 | 12% | 317 | 360 |
| So2 | 43 | 12% | 317 | 360 |
| maternal smoking (cotinine) | 10 | 3% | 350 | 360 |
| food insecurity | 29 | 8% | 331 | 360 |
| SRQ | 45 | 13% | 315 | 360 |
| IPV - emotional | 44 | 12% | 316 | 360 |
| IPV - physical | 44 | 12% | 316 | 360 |
| IPV - sexual | 44 | 12% | 316 | 360 |
| LEQ | 52 | 14% | 308 | 360 |
| EPDS | 44 | 12% | 316 | 360 |
| ASSIST - tobacco | 49 | 14% | 311 | 360 |
| ASSIST - alcohol | 49 | 14% | 311 | 360 |
| SES assets | 0 | 0% | 360 | 360 |
| Abbreviations: Particulate Matter (PM10); Carbon monoxide (CO); Nitrogen dioxide (NO2); Sulfur dioxide (SO2); Socioeconomic Status (SES); Self-Reporting Questionnaire (SRQ-20); Edinburgh Postnatal Depression Scale (EPDS); Life Experiences Questionnaire (LEQ); Intimate Partner Violence (IPV); Alcohol, Smoking, and Substance Involvement Screening Test (ASSIST) | | | | |

Table S3. Latent Class Mixed Effects Models (LCMM) model comparison statistics used to determine number of classes used in analysis. Bolded models were used in exposure-outcome analyses.

| Model Type | G | AIC | BIC | SABIC | entropy | %class1 | %class2 | %class3 | %class4 | %class5 |
| --- | --- | --- | --- | --- | --- | --- | --- | --- | --- | --- |
| Externalizing Problems | | | | | | | | | | |
| - | 1 | -45.14 | 17.08 | -33.68 | 1.00 | 100.00 |  |  |  |  |
| A | 2 | -50.47 | 31.20 | -35.43 | 0.56 | 44.04 | 55.96 |  |  |  |
| B | 2 | -59.27 | 18.51 | -44.94 | 0.61 | 48.20 | 51.80 |  |  |  |
| C | 2 | -57.78 | 23.88 | -42.74 | 0.61 | 52.63 | 47.37 |  |  |  |
| A | 3 | -79.80 | 21.31 | -61.18 | 0.82 | 26.04 | 52.08 | 21.88 |  |  |
| B | 3 | -79.80 | 21.31 | -61.18 | 0.82 | 52.08 | 26.04 | 21.88 |  |  |
| **C** | **3** | **-94.63** | **6.48** | **-76.00** | **0.85** | **48.48** | **22.71** | **28.81** |  |  |
| A | 4 | -69.81 | 50.75 | -47.60 | 0.86 | 51.80 | 26.04 | 22.16 | 0.00 |  |
| B | 4 | -95.08 | 25.48 | -72.87 | 0.85 | 39.34 | 11.63 | 23.27 | 25.76 |  |
| C | 4 | -96.11 | 24.45 | -73.90 | 0.80 | 28.25 | 23.27 | 26.87 | 21.61 |  |
| A | 5 | -104.14 | 35.86 | -78.35 | 0.83 | 24.65 | 27.70 | 14.40 | 23.55 | 9.70 |
| B | 5 | -104.14 | 35.86 | -78.35 | 0.83 | 9.70 | 23.55 | 24.65 | 27.70 | 14.40 |
| C | 5 | -127.27 | 12.72 | -101.49 | 0.87 | 8.03 | 21.05 | 21.05 | 28.81 | 21.05 |
| Internalizing Problems | | | | | | | | | | |
| - | 1 | 175.18 | 237.40 | 186.64 | 1.00 | 100.00 |  |  |  |  |
| A | 2 | 120.85 | 202.51 | 135.89 | 0.77 | 61.22 | 38.78 |  |  |  |
| B | 2 | 142.62 | 224.28 | 157.66 | 0.72 | 54.02 | 45.98 |  |  |  |
| C | 2 | 120.84 | 202.51 | 135.89 | 0.77 | 38.78 | 61.22 |  |  |  |
| A | 3 | 87.92 | 189.03 | 106.54 | 0.84 | 24.65 | 31.30 | 44.04 |  |  |
| B | 3 | 87.60 | 188.71 | 106.23 | 0.82 | 24.65 | 31.58 | 43.77 |  |  |
| **C** | **3** | **87.60** | **188.71** | **106.23** | **0.82** | **24.65** | **43.77** | **31.58** |  |  |
| A | 4 | 79.24 | 199.80 | 101.45 | 0.83 | 35.73 | 24.93 | 19.39 | 19.94 |  |
| B | 4 | 84.57 | 205.13 | 106.78 | 0.73 | 32.41 | 25.21 | 29.64 | 12.74 |  |
| C | 4 | 79.24 | 199.80 | 101.45 | 0.83 | 19.39 | 19.94 | 24.93 | 35.73 |  |
| A | 5 | 107.92 | 247.92 | 133.70 | 0.89 | 0.00 | 31.30 | 44.04 | 24.65 | 0.00 |
| B | 5 | 83.40 | 223.40 | 109.18 | 0.76 | 24.65 | 26.59 | 9.97 | 19.11 | 19.67 |
| C | 5 | 69.52 | 209.52 | 95.30 | 0.79 | 11.08 | 27.98 | 20.78 | 22.99 | 17.17 |
| Model Type A: Initial value generated from maximum likelihood estimates of a G=1 model; B: Initial value generated randomly from the asymptotic distribution of the estimates of the G=1 model; C: Initial value is found by a grid search run from 30 iterations from 100 random vectors.  Abbreviations: Number of classes (G), Akaike Information Criterion (AIC), Bayesian Information Criterion (BIC), sample-size-adjusted BIC (SABIC), Percentage of participants in class (%class). | | | | | | | | | | |

Table S4. Descriptive statistics (Median (IQR)) of indoor air pollutant and psychosocial factor exposures in Self-Organizing Map (SOM) exposure clusters.

|  | **SOM Cluster** | | | |
| --- | --- | --- | --- | --- |
|  | **1** | **2** | **3** | **4** |
| N (%) | 66 (18.3) | 147 (40.8) | 74 (20.6) | 73 (20.3) |
| PM10 µg/m3 (median [IQR]) | 29.06 [10.69, 58.50] | 38.63 [16.29, 65.70] | 39.39 [13.39, 68.55] | 39.70 [14.10, 73.82] |
| CO mg/m3 (median [IQR]) | 0.00 [0.00, 0.00] | 120.00 [0.00, 1050.00] | 0.00 [0.00, 120.00] | 0.00 [0.00, 60.00] |
| Benzene µg/m3 (median [IQR]) | 2.82 [0.95, 5.00] | 35.44 [12.66, 97.14] | 4.65 [2.55, 9.28] | 2.84 [1.30, 6.64] |
| Toluene µg/m3 (median [IQR]) | 10.67 [4.70, 21.48] | 53.72 [23.91, 255.05] | 18.69 [9.31, 46.87] | 13.57 [4.68, 24.37] |
| NO2 µg/m3 (median [IQR]) | 5.05 [2.09, 9.28] | 12.84 [7.28, 20.48] | 8.37 [4.24, 13.18] | 5.90 [3.44, 9.38] |
| SO2 µg/m3 (median [IQR]) | 0.00 [0.00, 0.26] | 0.00 [0.00, 0.64] | 0.00 [0.00, 0.26] | 0.00 [0.00, 0.00] |
| Urine Cotinine ng/ml (median [IQR]) | 16.50 [10.00, 50.88] | 21.50 [10.00, 47.00] | 500.00 [500.00, 500.00] | 431.00 [32.22, 500.00] |
| SES Asset Sum (median [IQR]) | 7.00 [6.00, 8.00] | 6.00 [4.00, 7.00] | 8.00 [7.00, 8.00] | 7.00 [5.00, 8.00] |
| Food Insecurity Total Score (median [IQR]) | 0.00 [0.00, 1.00] | 1.00 [0.00, 4.00] | 0.00 [0.00, 0.00] | 0.00 [0.00, 3.00] |
| SRQ-20 Total Score (median [IQR]) | 3.00 [1.00, 6.00] | 3.00 [1.00, 5.00] | 4.00 [2.00, 7.00] | 6.00 [4.00, 10.00] |
| EPDS Total Score (median [IQR]) | 8.00 [6.00, 12.00] | 10.00 [8.00, 13.00] | 8.00 [5.00, 12.00] | 11.00 [8.00, 15.00] |
| LEQ Total Score (median [IQR]) | 1.00 [0.00, 2.00] | 1.00 [0.00, 2.00] | 2.00 [1.00, 3.00] | 3.00 [1.00, 5.00] |
| Emotional IPV Score (median [IQR]) | 4.00 [4.00, 5.00] | 4.00 [4.00, 6.00] | 4.00 [4.00, 6.00] | 11.00 [9.00, 13.00] |
| Physical IPV Score (median [IQR]) | 5.00 [5.00, 6.00] | 5.00 [5.00, 7.00] | 5.50 [5.00, 6.75] | 11.50 [9.00, 15.00] |
| ASSIST Tobacco Score (median [IQR]) | 0.00 [0.00, 0.00] | 0.00 [0.00, 0.00] | 23.00 [17.00, 24.75] | 0.00 [0.00, 24.00] |
| ASSIST Alcohol Score (median [IQR]) | 0.00 [0.00, 0.00] | 0.00 [0.00, 0.00] | 0.00 [0.00, 10.00] | 0.00 [0.00, 14.25] |
| Abbreviations: Particulate Matter (PM10); Carbon monoxide (CO); Nitrogen dioxide (NO2); Sulfur dioxide (SO2); Socioeconomic Status (SES); Self-Reporting Questionnaire (SRQ-20); Edinburgh Postnatal Depression Scale (EPDS); Life Experiences Questionnaire (LEQ); Intimate Partner Violence (IPV); Alcohol, Smoking, and Substance Involvement Screening Test (ASSIST) | | | | |

Table S5. Pearson correlation coefficients among CBCL Total Problems, Externalizing Problems, and Internalizing Problems T-scores at each time point.

|  | 24 Months CBCL T-Score | | |
| --- | --- | --- | --- |
|  | Total Problems | Externalizing | Internalizing |
| Total Problems | 1 | 0.91 | 0.90 |
| Externalizing | 0.91 | 1 | 0.71 |
| Internalizing | 0.90 | 0.71 | 1 |
|  | 42 Months CBCL T-Score | | |
|  | Total Problems | Externalizing | Internalizing |
| Total Problems | 1 | 0.92 | 0.86 |
| Externalizing | 0.92 | 1 | 0.70 |
| Internalizing | 0.86 | 0.70 | 1 |
|  | 60 Months CBCL T-Score | | |
|  | Total Problems | Externalizing | Internalizing |
| Total Problems | 1 | 0.93 | 0.93 |
| Externalizing | 0.93 | 1 | 0.82 |
| Internalizing | 0.93 | 0.82 | 1 |

Table S6. Median (IQR) CBCL T-score at 24, 42, and 60 months for each CBCL trajectory.

|  |  | Median (IQR) CBCL T- Score | | |
| --- | --- | --- | --- | --- |
|  | N | 24 Months | 42 Months | 60 Months |
| Externalizing Problems | | | | |
| 1 (Medium) | 175 | 46.0 (15.0) | 40.0 (6.0) | 39.0 (12.0) |
| 2 (Low) | 80 | 43.0 (13.8) | 28.0 (0.0) | 38.0 (13.0) |
| 3 (High) | 105 | 50.0 (17.0) | 55.0 (8.0) | 43.0 (16.0) |
| Internalizing Problems | | | | |
| 1 (Increasing) | 89 | 51.0 (17.0) | 41.0 (16.0) | 61.0 (7.0) |
| 2 (Decreasing) | 158 | 43.0 (20.0) | 37.0 (12.00 | 29.0 (4.0) |
| 3 (Medium) | 113 | 47.0 (21.0) | 37.0 (18.0) | 43.0 (6.0) |

Table S7. Odds ratios and 95% CIs for individual exposure adjusted polytomous logistic regression models. Polytomous logistic regression models were adjusted for maternal HIV status, maternal age, and ancestry. Models using indoor air pollutant exposures were additionally adjusted for socioeconomic status, and principal components of psychosocial factors, and vice versa. Tables shows results from complete case models as well as multiple imputation (MI) models using 5 different random seeds (MI1 to MI5). MI4 models were presented in the main analysis.

|  | **Complete Case N** | **Complete Case** | **MI1** | **MI2** | **MI3** | **MI4 (Main results)** | **MI5** |
| --- | --- | --- | --- | --- | --- | --- | --- |
|  |  | **Externalizing Problems** | | | | | |
| PM10, high | 186 | 1.31 (0.95, 1.81) | 1.17 (0.95, 1.45) | **1.31 (1.05, 1.63)** | **1.32 (1.07, 1.64)** | **1.25 (1.01, 1.55)** | **1.25 (1.01, 1.55)** |
| PM10, medium |  | 1.11 (0.84, 1.47) | 1.04 (0.86, 1.24) | 1.1 (0.92, 1.33) | 1.13 (0.94, 1.35) | 1.15 (0.95, 1.38) | 1.09 (0.91, 1.3) |
| CO, high | 186 | 1.05 (0.89, 1.23) | 1 (0.89, 1.12) | 1.03 (0.92, 1.14) | 0.98 (0.88, 1.1) | 0.98 (0.87, 1.1) | 1.04 (0.93, 1.16) |
| CO, medium |  | 1.09 (0.93, 1.27) | 1.05 (0.95, 1.16) | 1.03 (0.93, 1.14) | 1.03 (0.93, 1.14) | 1.06 (0.96, 1.17) | 1.05 (0.95, 1.16) |
| Benzene, high | 186 | 1.05 (0.77, 1.43) | 0.94 (0.76, 1.16) | 1.01 (0.8, 1.27) | 1.04 (0.84, 1.29) | 1 (0.81, 1.25) | 0.94 (0.75, 1.17) |
| Benzene, medium |  | 0.99 (0.74, 1.32) | 0.84 (0.68, 1.02) | 0.9 (0.73, 1.12) | 0.94 (0.76, 1.15) | 0.91 (0.74, 1.11) | 0.94 (0.77, 1.16) |
| Toluene, high | 186 | 0.86 (0.66, 1.13) | 0.91 (0.75, 1.1) | 0.94 (0.78, 1.14) | 0.97 (0.81, 1.17) | 0.96 (0.79, 1.16) | 0.92 (0.75, 1.11) |
| Toluene, medium |  | 0.97 (0.77, 1.24) | 0.99 (0.83, 1.17) | 0.97 (0.82, 1.15) | 0.97 (0.82, 1.15) | 0.99 (0.83, 1.17) | 1.03 (0.86, 1.22) |
| NO2, high | 186 | 0.81 (0.53, 1.24) | 0.81 (0.6, 1.1) | 0.92 (0.69, 1.22) | 0.88 (0.65, 1.19) | 0.82 (0.6, 1.11) | 0.84 (0.62, 1.13) |
| NO2, medium |  | 0.93 (0.63, 1.38) | 0.97 (0.74, 1.27) | 0.97 (0.75, 1.26) | 0.92 (0.71, 1.21) | 0.89 (0.68, 1.16) | 0.93 (0.71, 1.21) |
| SO2, high | 186 | 0.73 (0.39, 1.37) | 0.85 (0.52, 1.38) | 0.75 (0.45, 1.23) | 0.84 (0.52, 1.36) | 0.75 (0.47, 1.19) | 0.81 (0.49, 1.33) |
| SO2, medium |  | 0.86 (0.51, 1.43) | 1.01 (0.68, 1.49) | 0.97 (0.67, 1.41) | 1.14 (0.78, 1.66) | 0.92 (0.64, 1.32) | 1 (0.68, 1.48) |
| Cotinine, high | 186 | 0.99 (0.63, 1.55) | 0.96 (0.74, 1.24) | 0.99 (0.76, 1.28) | 1 (0.76, 1.3) | 0.98 (0.75, 1.28) | 1.08 (0.84, 1.39) |
| Cotinine, medium |  | 1.2 (0.79, 1.83) | 1.16 (0.92, 1.47) | 1.09 (0.86, 1.38) | 1.15 (0.91, 1.47) | 1.17 (0.92, 1.49) | 1.21 (0.96, 1.52) |
| Food Insecurity, high | 184 | 0.76 (0.35, 1.66) | 0.86 (0.52, 1.45) | 0.68 (0.4, 1.15) | 0.8 (0.48, 1.33) | 0.81 (0.48, 1.36) | 0.87 (0.52, 1.46) |
| Food Insecurity, medium |  | 0.8 (0.4, 1.6) | 0.7 (0.44, 1.11) | 0.68 (0.43, 1.08) | 0.74 (0.47, 1.16) | 0.77 (0.49, 1.21) | 0.79 (0.5, 1.24) |
| SQR, high | 186 | 1.37 (0.77, 2.42) | 1.27 (0.87, 1.84) | 1.27 (0.87, 1.85) | 1.28 (0.89, 1.84) | 1.3 (0.89, 1.9) | 1.2 (0.82, 1.75) |
| SRQ, medium |  | 0.95 (0.57, 1.58) | 1.05 (0.75, 1.47) | 1.01 (0.72, 1.4) | 1.01 (0.73, 1.4) | 0.98 (0.7, 1.37) | 1.02 (0.72, 1.42) |
| IPV Emotional, high | 186 | 1.61 (0.44, 5.88) | 1.14 (0.48, 2.7) | 1.2 (0.5, 2.87) | 1.47 (0.61, 3.56) | 1.27 (0.52, 3.08) | 0.95 (0.4, 2.26) |
| IPV Emotional, medium |  | 1.41 (0.41, 4.9) | 1.09 (0.48, 2.47) | 1.23 (0.55, 2.74) | 1.3 (0.57, 2.99) | 1.47 (0.65, 3.35) | 1.2 (0.54, 2.67) |
| IPV Physical, high | 186 | 2.51 (0.52, 12.17) | 1.18 (0.43, 3.2) | 1.41 (0.51, 3.88) | 1.87 (0.68, 5.15) | 1.29 (0.47, 3.56) | 1.24 (0.47, 3.31) |
| IPV Physical, medium |  | 3.56 (0.78, 16.15) | 1.94 (0.78, 4.83) | 2.46 (0.98, 6.2) | **2.65 (1.03, 6.82)** | 2.47 (0.98, 6.24) | 1.92 (0.78, 4.72) |
| LEQ, high | 181 | 1.11 (0.57, 2.16) | 1.02 (0.65, 1.6) | 0.78 (0.49, 1.24) | 1.14 (0.72, 1.8) | 0.99 (0.63, 1.56) | 1.08 (0.68, 1.7) |
| LEQ, medium |  | 1.21 (0.65, 2.26) | 1.12 (0.74, 1.7) | 0.84 (0.55, 1.28) | 1.11 (0.73, 1.71) | 1.07 (0.7, 1.63) | 1.1 (0.72, 1.68) |
| EPDS, high | 186 | 0.82 (0.4, 1.68) | 0.96 (0.63, 1.47) | 1.03 (0.69, 1.54) | 0.98 (0.65, 1.47) | 0.96 (0.62, 1.47) | 1.01 (0.65, 1.57) |
| EPDS, medium |  | 0.52 (0.26, 1.03) | 0.67 (0.46, 0.99) | 0.82 (0.57, 1.19) | 0.87 (0.6, 1.28) | 0.7 (0.47, 1.04) | 0.7 (0.47, 1.05) |
| ASSIST Tobacco, high | 181 | 1.05 (0.66, 1.65) | 1.19 (0.91, 1.56) | 1.13 (0.89, 1.44) | 1.11 (0.86, 1.42) | 1.13 (0.87, 1.47) | 1.1 (0.85, 1.42) |
| ASSIST Tobacco, medium |  | 0.94 (0.62, 1.45) | 1.18 (0.91, 1.52) | 1.23 (0.98, 1.55) | 1.23 (0.97, 1.56) | 1.18 (0.92, 1.52) | 1.2 (0.94, 1.53) |
| ASSIST Alcohol, high | 181 | 0.84 (0.55, 1.29) | 1.15 (0.86, 1.54) | 1.16 (0.87, 1.55) | 1.16 (0.86, 1.56) | 1.25 (0.93, 1.69) | 1.07 (0.82, 1.4) |
| ASSIST Alcohol, medium |  | 0.71 (0.47, 1.09) | 0.98 (0.73, 1.31) | 1 (0.75, 1.33) | 1.08 (0.81, 1.45) | 1.07 (0.8, 1.45) | 0.88 (0.67, 1.15) |
| SES, high | 186 | 1.32 (0.27, 6.51) | 0.62 (0.2, 1.92) | 0.53 (0.17, 1.66) | 0.63 (0.21, 1.93) | 0.58 (0.19, 1.82) | 0.57 (0.18, 1.77) |
| SES, medium |  | 0.78 (0.18, 3.32) | 0.51 (0.18, 1.41) | 0.53 (0.19, 1.46) | 0.56 (0.21, 1.54) | 0.49 (0.17, 1.36) | 0.51 (0.19, 1.42) |
|  |  | **Internalizing Problems** | | | | | |
| PM10, increasing | 186 | 1.24 (0.92, 1.66) | 1.13 (0.93, 1.39) | 1.21 (0.98, 1.5) | **1.24 (1.01, 1.53)** | 1.22 (1, 1.5) | 1.13 (0.92, 1.38) |
| PM10, medium |  | 0.93 (0.73, 1.18) | 0.9 (0.76, 1.06) | 0.91 (0.77, 1.08) | 0.92 (0.78, 1.08) | 0.98 (0.83, 1.16) | 0.94 (0.79, 1.11) |
| CO, increasing | 186 | 1.04 (0.91, 1.18) | 1.05 (0.96, 1.16) | 1.04 (0.95, 1.15) | 1.05 (0.95, 1.16) | 1.05 (0.95, 1.16) | 1.05 (0.95, 1.15) |
| CO, medium |  | 1.05 (0.92, 1.19) | 1.02 (0.92, 1.12) | 1 (0.91, 1.1) | 1.07 (0.97, 1.17) | 1.04 (0.95, 1.15) | 1.04 (0.95, 1.14) |
| Benzene, increasing | 186 | 1.06 (0.82, 1.37) | 1.08 (0.88, 1.31) | 1.15 (0.93, 1.41) | 1.16 (0.94, 1.42) | **1.24 (1.02, 1.51)** | 1.09 (0.88, 1.33) |
| Benzene, medium |  | 0.96 (0.74, 1.23) | 1.06 (0.88, 1.28) | 1.09 (0.89, 1.33) | 1.15 (0.95, 1.38) | 1.09 (0.9, 1.32) | 1.11 (0.91, 1.34) |
| Toluene, increasing | 186 | 1.1 (0.88, 1.37) | 1.14 (0.97, 1.36) | **1.21 (1.02, 1.43)** | 1.14 (0.97, 1.35) | **1.21 (1.02, 1.44)** | 1.18 (0.99, 1.4) |
| Toluene, medium |  | 1.01 (0.81, 1.25) | 1.13 (0.96, 1.31) | 1.16 (0.99, 1.36) | 1.12 (0.96, 1.31) | 1.08 (0.92, 1.27) | 1.16 (0.99, 1.36) |
| NO2, increasing | 186 | 0.99 (0.7, 1.42) | 0.95 (0.73, 1.25) | 0.94 (0.73, 1.22) | 0.97 (0.74, 1.26) | 1.02 (0.78, 1.33) | 1.02 (0.78, 1.32) |
| NO2, medium |  | 0.83 (0.6, 1.16) | 0.84 (0.66, 1.07) | 0.84 (0.67, 1.07) | 0.83 (0.65, 1.06) | 0.84 (0.66, 1.07) | 0.86 (0.68, 1.09) |
| SO2, increasing | 186 | 0.75 (0.42, 1.36) | 0.82 (0.53, 1.28) | 0.77 (0.49, 1.21) | 0.82 (0.53, 1.29) | 0.78 (0.49, 1.23) | 0.84 (0.54, 1.33) |
| SO2, medium |  | 0.91 (0.59, 1.41) | 0.94 (0.66, 1.32) | 0.93 (0.66, 1.29) | 1.15 (0.86, 1.53) | 1.07 (0.78, 1.47) | 1.02 (0.73, 1.42) |
| Cotinine, increasing | 186 | 1.22 (0.84, 1.77) | 0.98 (0.78, 1.24) | 1.01 (0.8, 1.27) | 0.94 (0.74, 1.2) | 0.97 (0.77, 1.24) | 1.05 (0.84, 1.32) |
| Cotinine, medium |  | 0.98 (0.69, 1.4) | 0.91 (0.73, 1.13) | 0.92 (0.74, 1.14) | 0.87 (0.7, 1.09) | 0.93 (0.75, 1.16) | 0.91 (0.74, 1.13) |
| Food Insecurity, increasing | 184 | 1.49 (0.76, 2.91) | 1.3 (0.81, 2.08) | 1.08 (0.67, 1.74) | 1.1 (0.69, 1.75) | 1.11 (0.69, 1.77) | 1.19 (0.75, 1.89) |
| Food Insecurity, medium |  | 1.58 (0.85, 2.94) | 1.4 (0.92, 2.14) | 1.21 (0.8, 1.84) | 1.24 (0.82, 1.87) | 1.26 (0.84, 1.91) | 1.2 (0.79, 1.82) |
| SQR, increasing | 186 | 1.23 (0.76, 1.99) | 1.3 (0.92, 1.82) | 1.26 (0.9, 1.77) | 1.36 (0.98, 1.91) | 1.17 (0.83, 1.64) | 1.4 (0.99, 1.98) |
| SRQ, medium |  | 1.38 (0.87, 2.18) | 1.28 (0.94, 1.74) | 1.29 (0.95, 1.75) | 1.28 (0.94, 1.73) | 1.28 (0.93, 1.75) | **1.38 (1.01, 1.9)** |
| IPV Emotional, increasing | 186 | 1.6 (0.5, 5.15) | 1.54 (0.69, 3.48) | 1.54 (0.69, 3.45) | 1.3 (0.57, 2.97) | 1.52 (0.67, 3.46) | 1.89 (0.85, 4.22) |
| IPV Emotional, medium |  | **4.74 (1.66, 13.55)** | **2.56 (1.21, 5.42)** | **2.34 (1.12, 4.87)** | **2.58 (1.22, 5.46)** | **2.66 (1.27, 5.57)** | **2.78 (1.32, 5.84)** |
| IPV Physical, increasing | 186 | 0.85 (0.24, 2.97) | 1.16 (0.47, 2.82) | 1.24 (0.52, 2.97) | 1.13 (0.46, 2.76) | 1.08 (0.44, 2.63) | 1.11 (0.46, 2.66) |
| IPV Physical, medium |  | 2.15 (0.74, 6.22) | 1.96 (0.88, 4.39) | 1.72 (0.78, 3.82) | 2 (0.9, 4.45) | 1.95 (0.89, 4.27) | 1.9 (0.87, 4.17) |
| LEQ, increasing | 181 | 1.46 (0.82, 2.59) | **1.51 (1, 2.27)** | 1.12 (0.74, 1.71) | 1.44 (0.94, 2.19) | 1.27 (0.84, 1.92) | 1.25 (0.82, 1.89) |
| LEQ, medium |  | 1.07 (0.63, 1.82) | 1.23 (0.84, 1.8) | 1.16 (0.79, 1.71) | 1.15 (0.78, 1.7) | 1.21 (0.82, 1.77) | 1 (0.68, 1.48) |
| EPDS, increasing | 186 | 1.59 (0.9, 2.81) | 1.24 (0.86, 1.8) | 1.27 (0.87, 1.84) | 1.47 (0.99, 2.17) | 1.39 (0.94, 2.06) | **1.63 (1.08, 2.46)** |
| EPDS, medium |  | 1.25 (0.77, 2.03) | 1.09 (0.78, 1.53) | 1.05 (0.76, 1.46) | 1.18 (0.84, 1.66) | 1.28 (0.89, 1.83) | 1.23 (0.86, 1.76) |
| ASSIST Tobacco, increasing | 181 | 1.06 (0.72, 1.55) | 1.01 (0.79, 1.29) | 1.02 (0.82, 1.27) | 1 (0.79, 1.26) | 0.97 (0.77, 1.23) | 0.95 (0.75, 1.2) |
| ASSIST Tobacco, medium |  | 1.14 (0.79, 1.64) | 1.25 (0.99, 1.58) | 1.09 (0.88, 1.34) | 1.17 (0.95, 1.46) | 1.14 (0.91, 1.44) | 1.15 (0.92, 1.43) |
| ASSIST Alcohol, increasing | 181 | 0.75 (0.5, 1.12) | 0.88 (0.66, 1.16) | 0.96 (0.73, 1.27) | 1 (0.77, 1.32) | 0.98 (0.75, 1.28) | 0.98 (0.75, 1.29) |
| ASSIST Alcohol, medium |  | 0.96 (0.67, 1.37) | 1.08 (0.84, 1.38) | 1.08 (0.84, 1.38) | 1.1 (0.85, 1.42) | 1.1 (0.86, 1.42) | 1.2 (0.95, 1.52) |
| SES, increasing | 186 | 0.4 (0.1, 1.61) | 0.51 (0.18, 1.41) | 0.53 (0.19, 1.46) | 0.56 (0.21, 1.54) | 0.49 (0.17, 1.36) | 0.51 (0.19, 1.42) |
| SES, medium |  | 0.61 (0.17, 2.15) | 0.62 (0.2, 1.92) | 0.53 (0.17, 1.66) | 0.63 (0.21, 1.93) | 0.58 (0.19, 1.82) | 0.57 (0.18, 1.77) |
| Abbreviations: Particulate Matter (PM10); Carbon monoxide (CO); Nitrogen dioxide (NO2); Sulfur dioxide (SO2); Socioeconomic Status (SES); Self-Reporting Questionnaire (SRQ-20); Edinburgh Postnatal Depression Scale (EPDS); Life Experiences Questionnaire (LEQ); Intimate Partner Violence (IPV); Alcohol, Smoking, and Substance Involvement Screening Test (ASSIST) | | | | | | | |

Table S8. Joint exposure models using PCA to create Principal Components (PCs) of indoor air pollutant and psychosocial factor variables. The first five or 16 PCs were used as a joint exposure variables in polytomous logistic regression modeling. OR and 95% CI from adjusted polytomous logistic regression models. Polytomous logistic regression models adjusted for maternal HIV status, maternal age, and ancestry.

|  | **OR (95% CI)** |
| --- | --- |
|  | **Externalizing Problems** |
| PC1, High | **1.25 (1.02, 1.54)** |
| PC1, Medium | **1.27 (1.04, 1.54)** |
| PC2, High | 0.93 (0.76, 1.15) |
| PC2, Medium | 0.91 (0.75, 1.09) |
| PC3, High | 1.21 (0.93, 1.57) |
| PC3, Medium | **1.33 (1.04, 1.71)** |
| PC4, High | 0.91 (0.69, 1.21) |
| PC4, Medium | 1 (0.78, 1.3) |
| PC5, High | 0.83 (0.63, 1.1) |
| PC5, Medium | 1.04 (0.81, 1.34) |
|  | **Internalizing Problems** |
| PC1, increasing | 1.06 (0.88, 1.28) |
| PC1, medium | 1.15 (0.97, 1.36) |
| PC2, increasing | **1.22 (1.01, 1.48)** |
| PC2, medium | 1.03 (0.86, 1.22) |
| PC3, increasing | 0.97 (0.77, 1.23) |
| PC3, medium | 0.96 (0.77, 1.19) |
| PC4, increasing | 0.87 (0.67, 1.12) |
| PC4, medium | 0.81 (0.64, 1.03) |
| PC5, increasing | 0.8 (0.62, 1.04) |
| PC5, medium | 0.96 (0.76, 1.21) |

Table S9. Joint exposure models using SOM clusters as a joint exposure variables, OR and 95% CIs from adjusted polytomous regression models. Polytomous logistic regression models adjusted for maternal HIV status, maternal age, and ancestry.

|  | **OR (95% CI)** |
| --- | --- |
|  | **Externalizing Problems** |
| Cluster 1, high | REF |
| Cluster 1, Medium | REF |
| Cluster 2, High | 1.08 (0.46, 2.53) |
| Cluster 2, Medium | 0.8 (0.38, 1.69) |
| Cluster 3, High | **2.67 (1.14, 6.27)** |
| Cluster 3, Medium | 1.88 (0.82, 4.28) |
| Cluster 4, High | 1.57 (0.64, 3.85) |
| Cluster 4, Medium | 1.67 (0.74, 3.76) |
|  | **Internalizing Problems** |
| Cluster 1, increasing | REF |
| Cluster 1, medium | REF |
| Cluster 2, increasing | 1.91 (0.89, 4.1) |
| Cluster 2, medium | 1.21 (0.59, 2.47) |
| Cluster 3, increasing | 1.81 (0.85, 3.82) |
| Cluster 3, medium | 1.18 (0.56, 2.47) |
| Cluster 4, increasing | 1.2 (0.52, 2.79) |
| Cluster 4, medium | 1.75 (0.86, 3.59) |
| SOM cluster exposures detailed in figure S4A and table S4. LCMM trajectories detailed in Figure 1 and table S8. | |

Table S10. Individual polytomous logistic regression models additionally adjusted for season of indoor air pollution measurement. Polytomous logistic regression models adjusted for maternal HIV status, maternal age, and ancestry. Multiple Imputation (MI) seed 4 results shown.

|  | **OR (95% CI)** | |
| --- | --- | --- |
|  | **MI4 Main results** | **MI4 with Season** |
|  | **Externalizing Problems** | |
| PM10, high | **1.25 (1.01, 1.55)** | **1.29 (1.02, 1.63)** |
| PM10, medium | 1.15 (0.95, 1.38) | 1.12 (0.92, 1.36) |
| CO, high | 0.98 (0.87, 1.1) | 1.01 (0.89, 1.16) |
| CO, medium | 1.06 (0.96, 1.17) | 1.06 (0.94, 1.19) |
| Benzene, high | 1 (0.81, 1.25) | 1 (0.77, 1.29) |
| Benzene, medium | 0.91 (0.74, 1.11) | 0.84 (0.67, 1.07) |
| Toluene, high | 0.96 (0.79, 1.16) | 0.94 (0.76, 1.18) |
| Toluene, medium | 0.99 (0.83, 1.17) | 0.92 (0.76, 1.11) |
| NO2, high | 0.82 (0.6, 1.11) | 0.87 (0.62, 1.20) |
| NO2, medium | 0.89 (0.68, 1.16) | 0.98 (0.74, 1.13) |
| SO2, high | 0.75 (0.47, 1.19) | 0.80 (0.46, 1.38) |
| SO2, medium | 0.92 (0.64, 1.32) | 1.09 (0.71, 1.67) |
|  | Internalizing Problems | |
| PM10, increasing | 1.22 (1, 1.5) | 1.13 (0.91, 1.40) |
| PM10, medium | 0.98 (0.83, 1.16) | 0.89 (0.74, 1.06) |
| CO, increasing | 1.05 (0.95, 1.16) | 1.07 (0.95, 1.19) |
| CO, medium | 1.04 (0.95, 1.15) | 1.05 (0.94, 1.17) |
| Benzene, increasing | **1.24 (1.02, 1.51)** | 1.18 (0.94, 1.48) |
| Benzene, medium | 1.09 (0.9, 1.32) | 1.08 (0.87, 1.35) |
| Toluene, increasing | **1.21 (1.02, 1.44)** | **1.25 (1.03, 1.51)** |
| Toluene, medium | 1.08 (0.92, 1.27) | 1.12 (0.93, 1.34) |
| NO2, increasing | 1.02 (0.78, 1.33) | 0.99 (0.75, 1.31) |
| NO2, medium | 0.84 (0.66, 1.07) | 0.82 (0.63, 1.07) |
| SO2, increasing | 0.78 (0.49, 1.23) | 0.79 (0.49, 1.27) |
| SO2, medium | 1.07 (0.78, 1.47) | 1 (0.70, 1.42) |
| Abbreviations: Particulate Matter (PM10); Carbon monoxide (CO); Nitrogen dioxide (NO2); Sulfur dioxide (SO2) | | |

**Supplemental Figures**

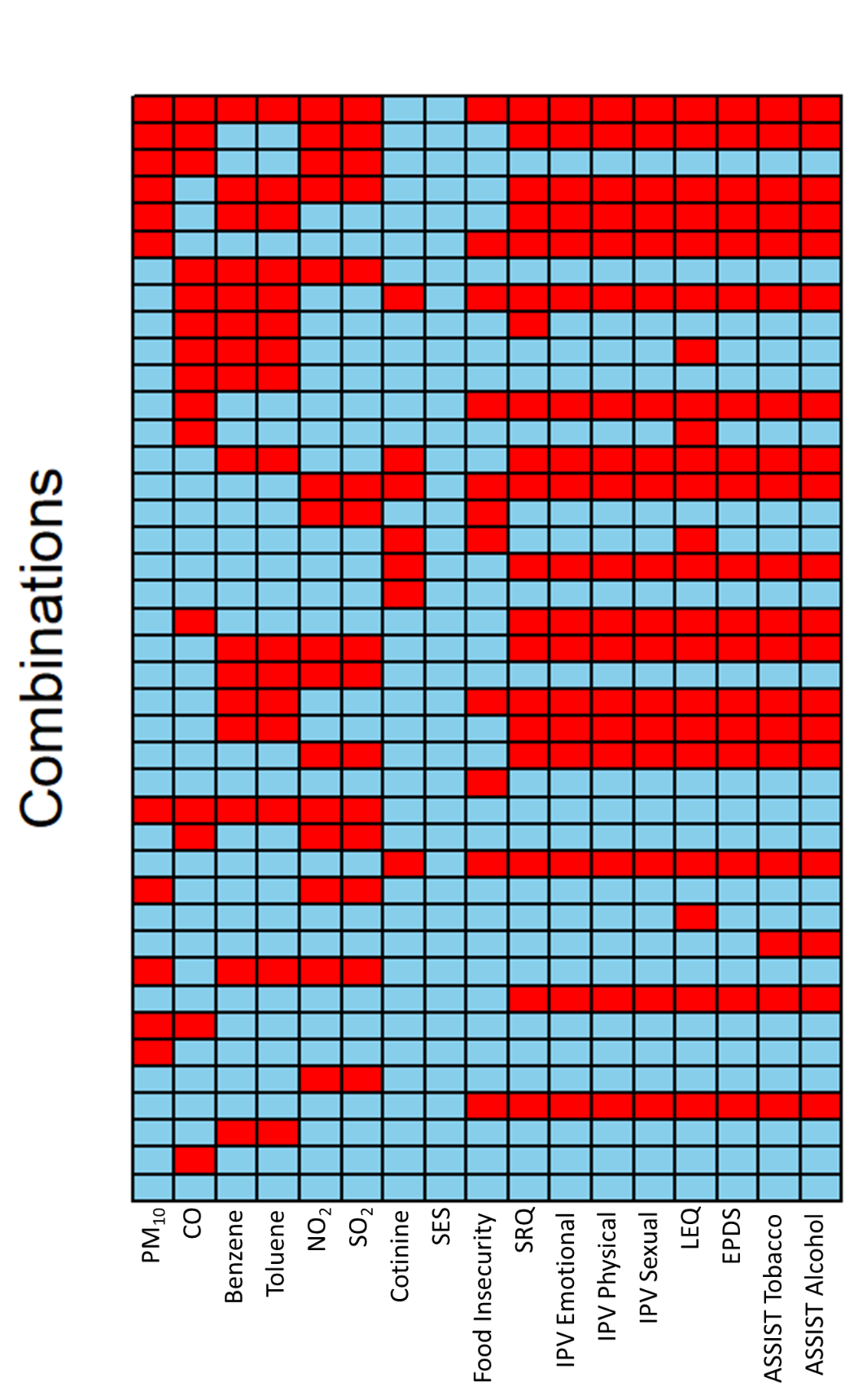

Figure S1. Combinations of missingness patterns of exposure variables. Each row is a missingness pattern where red indicates that variable is missing, and blue indicates not missing.

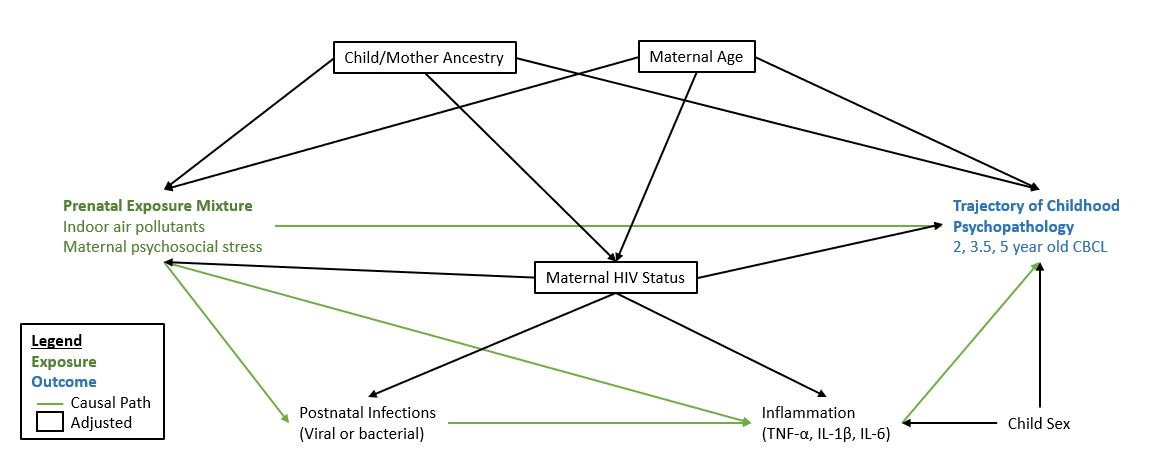

Figure S2. DAG of underlying causal pathways between prenatal exposure to indoor air pollutants and psychosocial factors including socioeconomic status, and trajectories of childhood psychopathology.

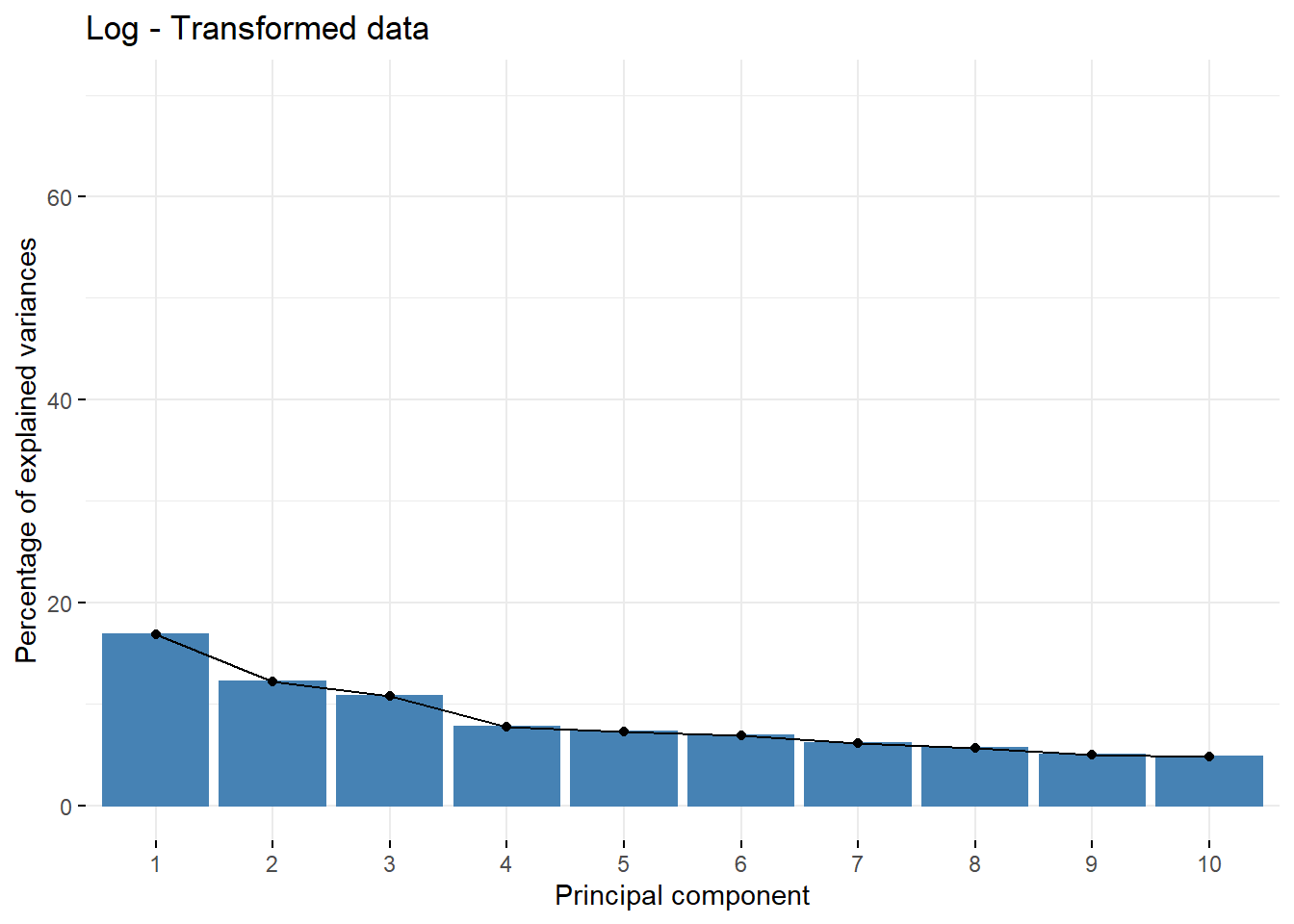

Figure S3. Scree (elbow) plot describing the proportion of variance explained by each principal component analysis of indoor air pollutant and psychosocial factor exposures.

A.

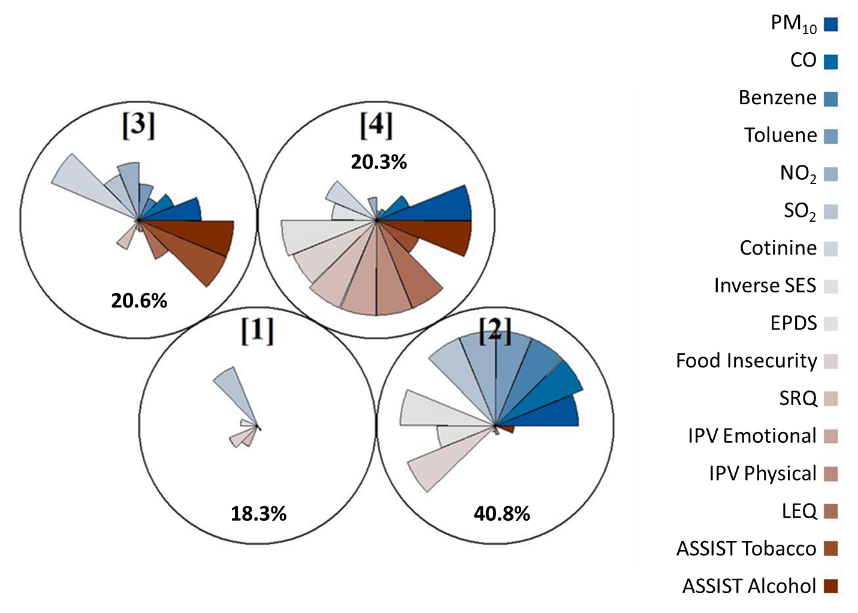

B.	C.

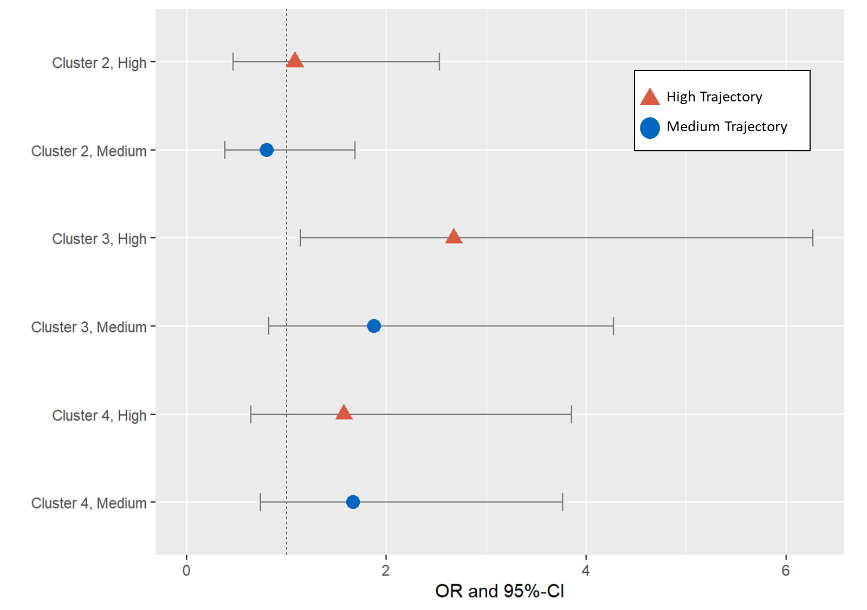

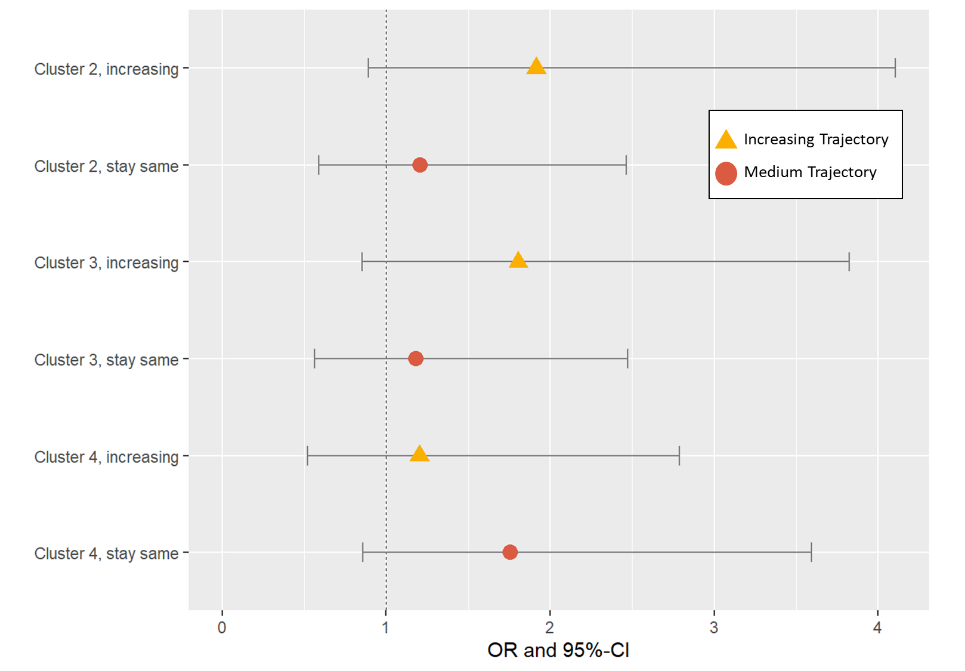

Figure S4. SOM analysis investigating associations between clusters of indoor air pollution and psychosocial factors on CBCL trajectories. A. SOM cluster star plot, slices represent median values of the mixture component, each circle is a SOM cluster. Blue slices represent indoor air pollutants while red slices represent psychosocial factors. B. Results from adjusted polytomous logistic regression model for Externalizing Problems trajectory, using cluster 1 as the reference group. Model adjusted for maternal age, maternal HIV status, and ancestry. C. Results from adjusted polytomous logistic regression model for Internalizing Problems trajectory, using cluster 1 as the reference group. Model adjusted for maternal age, maternal HIV status, and ancestry.
